# Predictors of adherence to public health instructions during the COVID-19 pandemic

**DOI:** 10.1101/2020.04.24.20076620

**Authors:** Yehuda Pollak, Haym Dayan, Rachel Shoham, Itai Berger

## Abstract

**Importance:** Identifying risk factors for adherence to public health instructions for the COVID-19 pandemic may be crucial for controlling the rate of transmission and the pandemic’s health and economic impacts.

**Objective:** To determine sociodemographic, health-related, risk-related, and instruction-related factors that predict non-adherence to instructions for the COVID-19 pandemic.

**Design:** Cross-sectional survey in Israel collected between March 28 and April 10, 2020.

**Setting:** Population-based study.

**Participants:** A convenience sample completed an online survey.

**Exposures:** Sociodemographic, health-related, risk-related, and instruction-related characteristics of the participants that have been linked to adherence to medical instructions.

**Main Outcome and Measure:** Non-adherence to instructions defined by a mean score of less than 4 on a 1 to 5 adherence scale consisting of 19 instruction items.

**Results:** Among 654 participants (413 [64.8%] female, age 40.14 [15.23] years), 28.7% were defined as non-adherents. Non-adherence was associated with male gender [adjusted odds ratio (aOR) = 1.54, CI 1.03– 2.31], not having children [aOR = 1.73, 1.13–2.65], smoking [aOR = 2.27, CI 1.42–3.62], high levels of ADHD symptoms [aOR = 1.55, CI 1.07–2.25], high levels of past risk-taking behavior [aOR = 1.41, CI 1.10–1.81], as well as by current high psychological distress [aOR = 1.51, CI 1.14–2.01], low perceived risk of COVID-19 [aOR = 1.52, CI 1.22–1.89], low exposure to the instructions [aOR = 1.45, CI 1.14–1.82], and low perceived efficacy of the instructions [aOR = 1.47, CI 1.16–1.85]. Adjusted OR of age, economic status, physical health status, and exposure to media did not reach the significance level.

**Conclusions and Relevance:** People with the above characteristics may have increased risk for non-adherence to public health instructions. There appears to be a need for setting out and communicating instructions to specifically targeted at-risk populations.

**Key Points:** *Question:* What factors are associated with non-adherence to public health instructions during COVID-19?

*Findings:* In a cross-sectional study of 654 Israeli participants, non-adherence to instructions was associated with male gender, not having children, smoking, high levels of attention-deficit/hyperactivity disorder (ADHD) symptoms, low level of pro-sociality, and high levels of past risk-taking behavior, as well as by current high psychological distress, high perceived risk of the COVID-19, high exposure to the instructions, and high perceived efficacy of the instructions.

*Meaning:* The findings suggest that in setting out and communicating public health instructions, policymakers should consider the above sociodemographic, health-related, risk-related, and instruction-related characteristics.

## Introduction

The novel coronavirus 2019 (COVID-19) outbreak has had an enormous global impact. In response to the rapidly spreading pandemic, states have introduced public health measures to limit community transmission of COVID-19. In Israel, between February 2 and March 17, 2020, the Ministry of Health gradually introduced a series of orders concerning home-isolation and quarantine, personal hygiene, restrictions on gathering and traveling, and social distancing.

The public health instructions’ efficacy in limiting the spread of the pandemic depends on public adherence to them. Despite the potentially harmful consequences for individuals and public health, non-adherence to the instructions (non-AtI) for the COVID-19 pandemic has been frequent.^1^ The objective of this study was to identify predictors of non-adherence. The hypothesized predictors were chosen based on the clinical literature regarding the risk factors for non-adherence to medical instructions, and more generally, for engagement in risk-taking behavior,^2–4^ and more generally, for engagement in risk-taking behavior.^5,6^ These include demographic/economic factors (i.e., young age, male gender, low religiousness, unemployment status, and low-income level), health and personality factors (i.e., background and current physical and mental health problems, attention-deficit/hyperactivity disorder (ADHD) symptoms, low levels of prosociality, past risk-taking behavior), perceptions regarding COVID-19 risk and efficacy of the instructions, and low exposure to the instructions.

## Methods

This study was approved by the ethics committee of the Seymour Fox School of Education at the Hebrew University of Jerusalem.

From March 28 to April 10, 2020, a convenience sample of 849 participants filled out an online survey. Participants were recruited by social media (WhatsApp groups, Twitter, Facebook). Of the total sample, 654 participants completed the AtI scale and were enrolled to the study. The dependent variable, non-AtI, was measured by 20 questions probing for adherence to each of the instructions released to the public by the Ministry of Health at that time. For each item, participants were asked to rate their adherence on a Likert scale, ranging from not at all (1) to very strictly (5) (see Table S1 in the supplementary materials). The mean score on the AtI scale was calculated for each participant. Non-adherence was defined by a mean response of <4 across the AtI scale.

To estimate the contribution of the predictors of non-AtI, the following tools were used A sociodemographic questionnaire included specific questions regarding each participant’s age, gender, marital status, number of children, level of education, ethnicity, religiousness, place of birth, level of income, and working status. Health and personality factors were measured by specific questions regarding regular number of hours of sleep, physical activity, smoking, presence of chronic illness, and a single-item self-rated health measure on a Likert scale of 1–10. Psychological distress was measured by the K6 scale,^7^ and ADHD symptoms were measured by the Adult ADHD Self-Reporting Scale (ASRS-v1.1).^8^ Pro-sociality was measured by the pro-social subscale of the adult version of the Strength and Difficulties Questionnaire (SDQ).^9^ Past risk-taking behavior was measured by the Adult Risk Taking Inventory.^10^ Perceptions regarding COVID-19 risk and the efficacy of instructions in reducing this risk were measured by scales created for the purpose of this study: the Perceived Risk of COVID-19 Scale and the Perceived Efficacy of the Ministry of Health Instructions Scale. For detailed description of the measures, see the supplementary materials.

Descriptive statistics present the number of respondents and the percentages for categorical variables, and means and standard deviations for continuous variables. Logistic regression analyses were used to calculate the associations between independent variables and the primary outcomes. Variables that appeared to be associated (p<0.10) in the unadjusted analyses were further adjusted for demographic factors (i.e., age, gender, having children, education, and ethnicity) using stepwise logistic regressions. Associations with a p-value <0.05 in the adjusted analyses were considered to be statistically significant.

## Results

The overall sample is described in Table 1. In summary, of the 654 respondents, 15.2% were 18 to 24 years or older, 8.5% were 65 years or older, 64.8% were women, 67.1% were married / in a relationship, 87.7% identified as Jewish, 57% identified as religious, 92.6% were born in Israel, 75.2% lived in cities, 71.6% had higher education, 32.2% earned less than the mean income, 32.3% earned more than the mean income, 9.7% were unemployed before the COVID-19 crisis, and 39.2% were unemployed due to the crisis. Descriptive statistics of the K6, ASRS, pro-sociality, perceived risk, perceived effectiveness, and perceived illness are presented in Table 2. The distribution of the ratings of adherence to each instruction is provided in Table S1 in the supplementary materials. The minority of the participants (28.7%) reported a mean level lower than 4 and were consequently defined as non-adherents.

**Table 1:**
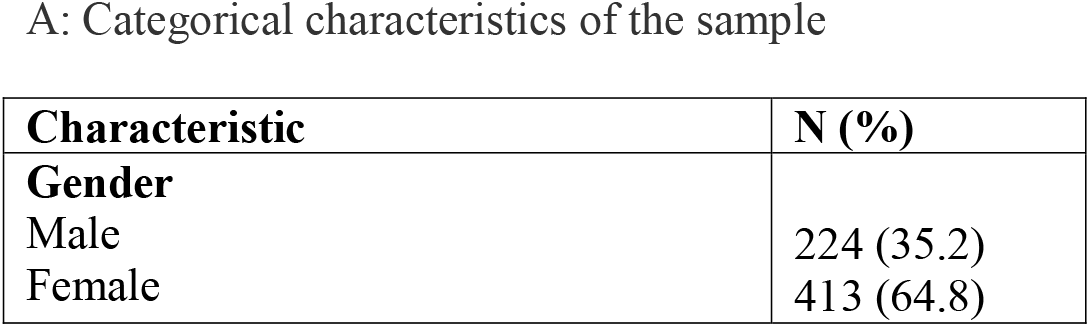
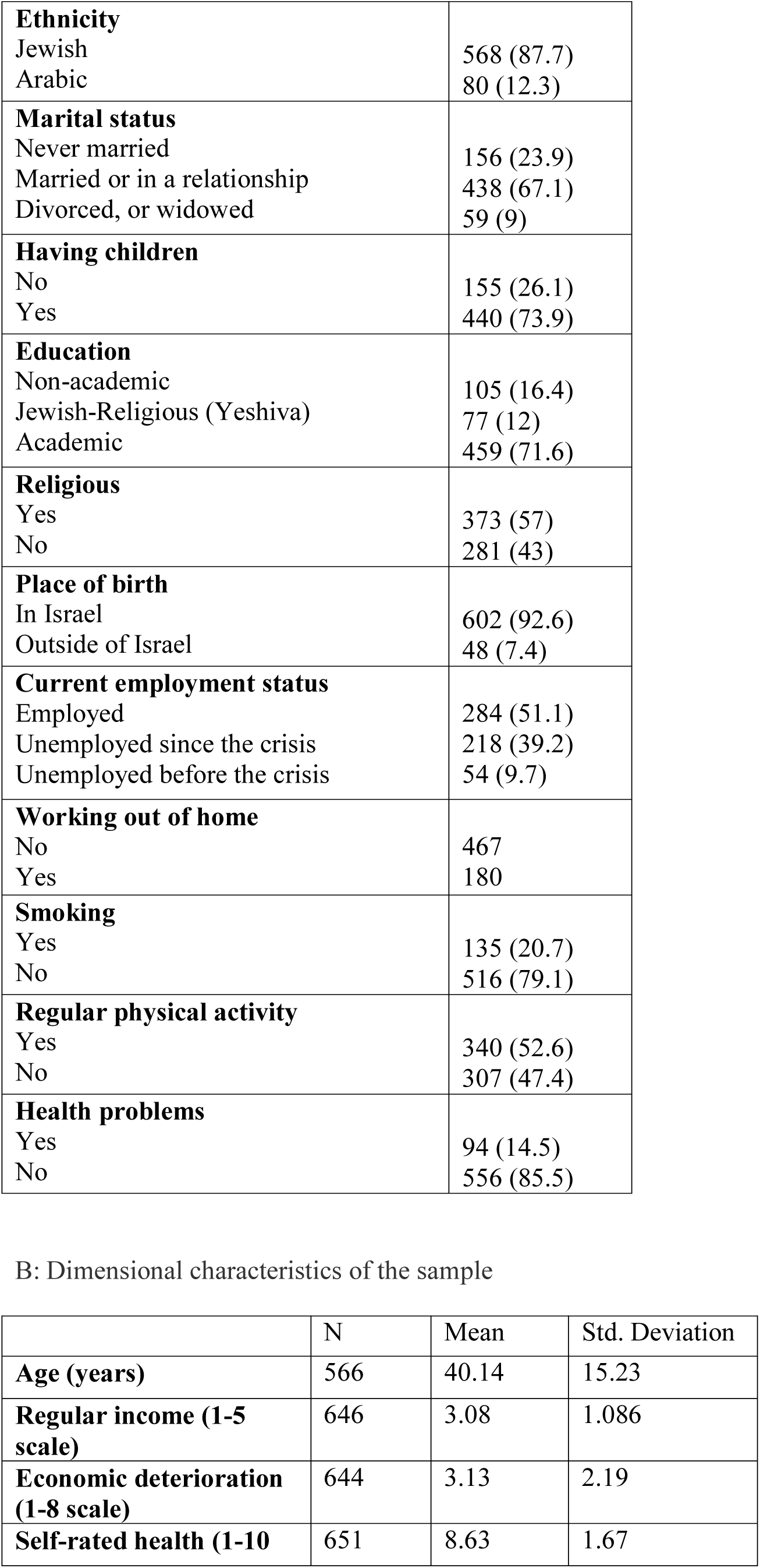
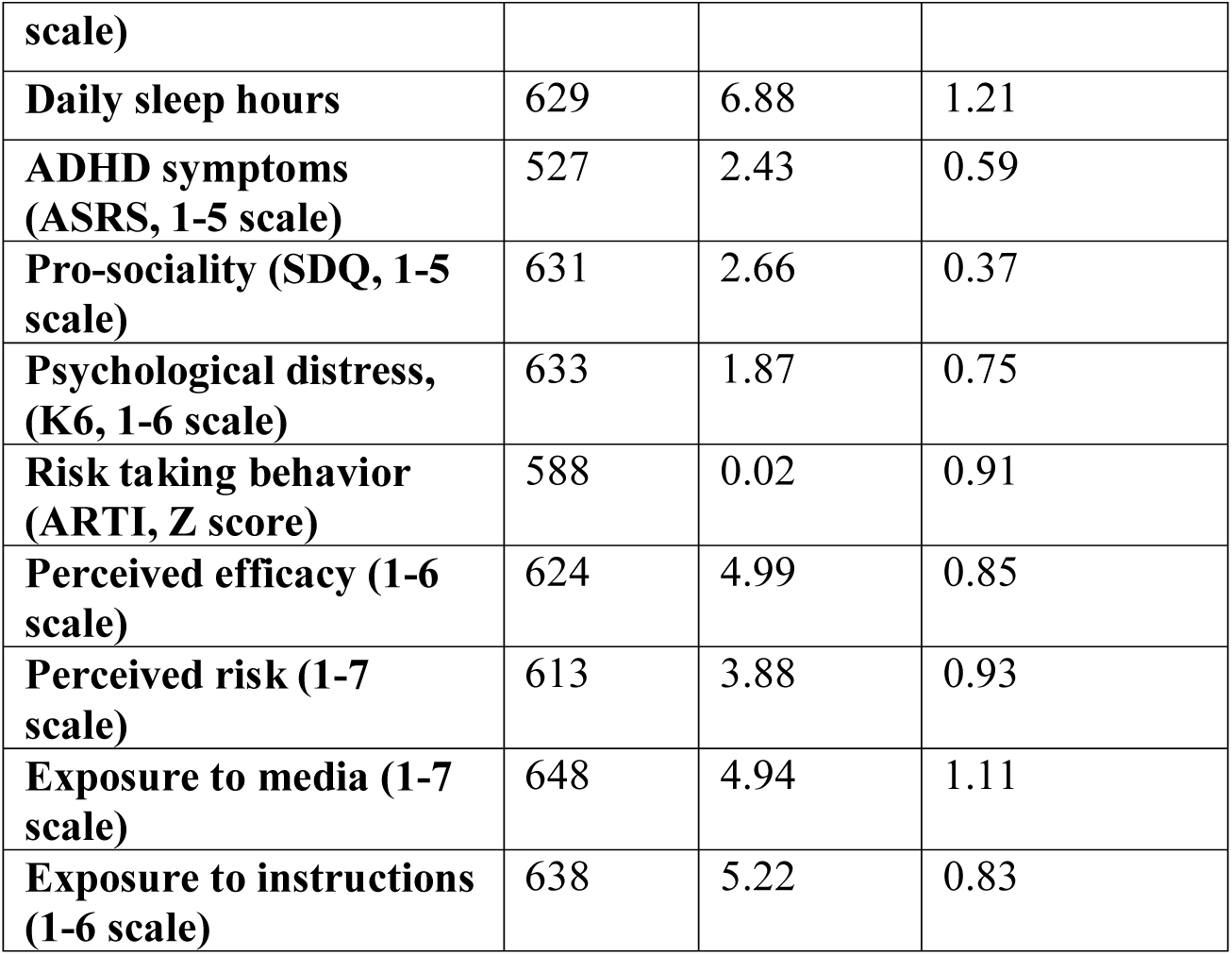
Sociodemographic, health-related, risk-related, and instruction-related categorical (A) and dimensional (B) characteristics of the sample

Table 3 presents the unadjusted and adjusted regression analysis results for non-AtI. The following background variables predicted non-adherence on adjusted analyses: male gender, not having children, smoking, high levels of ADHD symptoms, and high levels of past risk-taking behavior. Non-AtI was also predicted by the following current variables: high psychological distress, low perceived risk of COVID-19, low exposure to the instructions, and low perceived efficacy of the instructions.

**Table 3:** Non-adherence to the instructions of the Ministry of Health for the COVID-19 pandemic by a range of sociodemographic, health-related, risk-related, and instruction-related factors

|  | N (%) | Unadjusted OR | Adjusted OR |
| --- | --- | --- | --- |
| **Gender** |  |  |  |
| Male | 83 (37.1) | Ref | Ref |
| Female | 101 (24.5) | **1.81 (1.28-2.59)\*\*** | **1.54 (1.03-2.31)\*** |
| **Age (years)** | 566 | 0.99 (0.98-1.00) | 1.00 (.99-1.02) |
| **Ethnicity** |  |  |  |
| Jewish | 168 (29.6) | Ref | Ref |
| Arabic | 16 (20.0) | 1.68 (0.94-2.99) | 1.97 (0.92-4.20) |
| **Marital status** |  |  |  |
| Divorced, or widowed | 15 (25.4) | Ref |  |
| Never married | 53 (34.0) | 1.09 (0.59-2.04) |  |
| Married or in a relationship | 119 (27.2) | 1.51 (0.77-2.96) |  |
| **Having children** |  |  |  |
| Yes | 117 (26.6) | Ref | Ref |
| No | 53 (34.2) | 1.43 (0.97-2.13) | **1.73 (1.13-2.65)\*** |
| **Education** |  |  |  |
| Academic | 124 (27.0) | Ref | Ref |
| Jewish-Religious (Yeshiva) | 30 (39.0) | **1.72 (1.04-2.85)\*** | 1.25 (0.72-2.16) |
| Secondary school | 32 (30.5) | 1.18 (0.75-1.88) | 1.19 (0.60-2.38) |
| **Religious** |  |  |  |
| Ultraorthodox | 66 (27.0) | Ref |  |
| Orthodox | 31 (26.3) | 0.96 (0.58-1.58) |  |
| Traditional | 18 (27.3) | 1.01 (0.55-1.86) |  |
| Non-religious | 71 (33.0) | 1.33 (0.89-1.99) |  |
| **Place of birth** |  |  |  |
| Outside of Israel | 12 (25.0) | Ref |  |
| In Israel | 175 (29.1) | 1.23 (0.63-2.42) |  |
| **Level of income (1-5 scale)** | 646 | 0.95 (0.81-1.11) |  |
| **Current employment status** |  |  |  |
| Employed | 79 (27.8) | Ref |  |
| Unemployed since the crisis | 70 (32.1) | 1.23 (0.84-1.80) |  |
| Unemployed before the crisis | 16 (29.6) | 1.09 (0.58-2.07) |  |
| **Economic deterioration (1-7 scale)** | 644 | 0.97 (0.90-1.05) |  |
| **Working out of home** |  |  |  |
| No | 125 (26.8) | Ref |  |
| Yes | 62 (34.4) | 1.43 (0.99-2.08) | 1.30 (0.84-2.00) |
| **Daily sleep hours** | 629 | 1.12 (0.97-1.29) |  |
| **Smoking** |  |  |  |
| No | 126 (24.4) | Ref |  |
| Yes | 61 (45.2) | **2.55 (1.72- 3.78)\*\*\*** | **2.27 (1.42-3.62)\*\*** |
| **Regular physical activity** |  |  |  |
| Yes | 91 (26.8) | Ref |  |
| No | 95 (30.9) | 0.82 (0.58-1.15) |  |
| **Health problems** |  |  |  |
| Yes | 27 (28.7) | Ref |  |
| No | 160 (28.8) | 0.99 (0.62-1.62) |  |
| **Self-rated health (1-10 scale)** | 651 | 0.99 (0.89-1.09) |  |
| **ADHD symptoms (ASRS, 1-5 scale)** | 527 | **1.65 (1.20-2.27)\*\*** | **1.55 (1.07-2.25)\*** |
| **Pro-sociality (SDQ, 1-5 scale)** | 631 | **0.57 (0.36-0.90)\*\*** | 0.81 (0.46-1.40) |
| **Psychological distress, (K6, 1-6 scale)** | 633 | **1.37 (1.10-1.71)\*\*** | **1.51 (1.14-2.01)\*\*** |
| **Risk taking behavior (ARTI, Z score)** | 588 | **1.40 (1.15-1.72)\*\*** | **1.41 (1.10-1.81)\*\*** |
| **Perceived efficacy (1-6 scale)** | 624 | **0.65 (0.53-0.80)\*\*\*** | **0.68 (0.54-0.86)\*\*** |
| **Perceived risk (1-7 scale)** | 613 | **0.65 (0.54-0.78)\*\*\*** | **0.66 (0.53-0.82)\*\*\*** |
| **Exposure to media (1-7 scale)** | 648 | 0.92 (0.79-1.07) |  |
| **Exposure to instructions (1-6 scale)** | 638 | **0.66 (0.53-0.80)\*\*\*** | **0.69 (0.55-0.88)\*\*** |

*Note* \* $p < .05$ , \*\* $p < .01$ , \*\*\* $p < .001$

Note *p < .05, **p < .01, ***p < .001

## Discussion

In the absence of a vaccine and treatments, high adherence to public health instructions is essential for reducing transmission and the impact of COVID-19. Our study suggests that non-adherence to public health instructions for COVID-19 can be predicted by several demographic, health-related, risk-related, and instruction-related variables. Specifically, background male gender, not having children, high levels of ADHD symptoms, smoking, and high levels of past risk-taking behavior predicted non-adherence. As these factors preceded the outbreak and the public health instructions, they can be considered as causes or risk factors. Non-adherence was also predicted by current high distress levels, low exposure to the instructions, and low perceptions regarding the risk of COVID-19 and the efficacy of the instructions. As the latter factors coincided with the outbreak and the instructions, their causal relation cannot be determined^11^ and should be further studied by a longitudinal research design.

The findings of a link between non-AtI and being male and between non-AtI and low perceived risk are in line with other reports of males and non-anxious persons being less engaged in social distancing measures for coping with COVID-19 infection,^1,12^ as well as with reports concerning other epidemics.^13,14^ Other studies provided evidence for non-AtI among the most economically disadvantaged in society, which was not replicated in our sample, possibly due to differences in the operationalization of non-adherence and economic position. Our study, driven by findings of factors associated with risk-taking behavior in general,^5^ highlights the role of two background characteristics in predicting non-AtI – ADHD symptoms and previous engagement in risk-taking behavior – as well as a link with risk perception.

Our study has some strengths, including the use of scales, rather than single-item questions. The size of the sample, and the focus on the Israeli public and on the first weeks after the release of the instructions might limit the generalization of the findings to other places and times. Future studies should scrutinize adherence pattern variations throughout the epidemic. Further research is warranted to effectively design public health messaging targeted at populations at risk of non-adherence.

## Data Availability

The dataset analyzed during the current study will be available from the corresponding author on request.

